# Frailty-based prognostication of clinical outcomes in geriatric burn patients: a retrospective study

**DOI:** 10.64898/2026.05.12.26353034

**Authors:** Joanna H. Lee, Monica Jinsi, Michael Feldman, Sarah Hobgood

## Abstract

Burn trauma disproportionately impacts older adults, yet existing burn severity models emphasize age, total body surface area (TBSA), and inhalation injury without accounting for geriatric-specific vulnerabilities such as frailty. We conducted a retrospective cohort study of 326 geriatric patients admitted with burn injuries between 2020 and 2024 to evaluate how TBSA, burn location, inhalation injury, renal insufficiency, comorbidities, and functional dependence in activities of daily living (ADLs) and instrumental activities of daily living (IADLs) affect in-hospital mortality and discharge disposition. Based on multivariable logistic regression and chi-square analyses, TBSA, as expected, emerged as the strongest predictor across models. Each 1% increase in TBSA was associated with a 7% increase in the odds of in-hospital mortality (*p*=0.006) and a 12 to 19% increase in odds of post-acute care placement (*p*<0.001). Inhalation injury and renal insufficiency were also independently associated with increased odds of both mortality and post-acute care disposition, whereas respiratory comorbidity predicted mortality alone. Functional status demonstrated outcome-specific prognostic value: ADL dependence predicted mortality, while IADL dependence predicted discharge disposition. Patients with some ADL dependence had five-fold higher odds of in-hospital mortality (*p*=0.011), while some (OR=2.48, *p*= 0.039) and full IADL dependence (OR=2.61, *p*=0.025) were associated with higher odds of post-acute care placement. Integrating structured functional assessments that distinguish basic from instrumental limitations alongside established burn severity metrics may enhance prognostication and guide individualized care planning for older adults with burn injuries.

## INTRODUCTION

Burn injuries impose a major global health challenge, affecting a substantial and growing number of individuals with an increasing burden in part by growing population.^1^ In the United States, the Center for Disease Control and Prevention (CDC) data reported 398,000 fire-or burn-related injuries and an additional 252,000 contact burns from hot objects or substances in 2021.^2^

Older adults represent a rapidly expanding and uniquely vulnerable subset of the burn population. National epidemiologic analyses consistently show that geriatric patients (aged ≥65 years) experience higher mortality, increased complications, and worse functional outcomes following burn trauma compared with younger adults.^3–4^ This reflects both age-related physiological vulnerability and greater comorbidity burden. U.S. National Inpatient Sample show that in-hospital mortality increases with age, reaching nearly 10% among patients aged 85 and older.^5^ Moreover, longitudinal studies demonstrate that mortality remains elevated well beyond the index hospitalization for burn injuries: approximately 12% of elderly patients die during the initial admission, an additional 8% die within the first year after discharge, and up to 26% of discharged patients die within five years.^6^ As the U.S. population continues to age, the clinical and economic impact of burn injuries in older adults is expected to intensify. These findings underscore the need for improved risk stratification and prognostic assessment of geriatric burn patients.

Current burn prognostic models, such as the Baux score, Fatality by Longevity, APACHE II score, Measured Extent of burn, and Sex (FLAMES) score, and Abbreviated Burn Severity Index (ABSI), primarily incorporate age, TBSA, burn depth or location, and inhalation injury.^7^ Although these indices are widely used, they do not account for geriatric-specific risk factors such as frailty, baseline functional dependence, or chronic disease burden, despite growing evidence that these factors meaningfully shape outcomes in older burn patients.

Meanwhile, frailty, defined as decreased physiologic reserve and increased vulnerability to stressors, has emerged as an important predictor of outcomes in older adults with burn injuries, with multiple studies demonstrating its association with mortality, morbidity, and discharge disposition. Higher admission frailty scores are consistently linked to increased 30-day mortality, greater healthcare utilization, and a higher likelihood of discharge to skilled nursing facilities or rehabilitation, even after adjusting for age, total body surface area (TBSA), and inhalation injury.^8–10^ The Canadian Study of Health and Aging Clinical Frailty Scale (CFS) has shown predictive validity for in-hospital mortality, acute respiratory failure, and adverse discharge destinations.^9–11^ However, frailty assessments in burn care remain under-validated, and existing studies conceptualize frailty as a unidimensional exposure that aggregates functional dependence, comorbidity burden, cognitive impairment, and social vulnerability into a single clinician-scored metric.^12^ This approach may obscure which specific geriatric vulnerabilities most strongly influence outcomes and introduce subjectivity that can limit reproducibility and bedside applicability.

These limitations underscore the need for prognostic tools that more precisely integrate geriatric vulnerability into burn outcome prediction. To address this gap, our preliminary retrospective cohort study examined frailty domains alongside key burn characteristics, demonstrating that burn severity, functional dependence, and renal insufficiency were key predictors of hospital outcomes.^13^ Of note, this study was constrained by a modest sample size and limited generalizability.

The present study leverages a larger patient cohort and a more comprehensive set of clinical variables, evaluating functional dependence, comorbid conditions, and physiologic vulnerabilities in conjunction with traditional burn metrics such as TBSA, burn location, inhalation injury, and renal insufficiency. By analyzing how these domains collectively influence mortality, length of stay, and discharge disposition, this study aims to clarify how these distinct frailty domains relate to outcomes and improve risk stratification in older adults with burn injuries and inform the development of a burn-specific frailty prognostic tool, the B-FRAIL scale.

## METHODS

### Study design

We conducted a retrospective cohort study of geriatric patients with burn injuries admitted to a nationally-verified Level I burn center between January 1, 2020 and December 31, 2024. Patient data meeting inclusion criteria were obtained from the institutional burn registry, which prospectively captures demographic, clinical, and injury-related information for all admitted burn patients. These data were supplemented by detailed chart review of electronic medical records (EMR).

### Study population

Patients were eligible for inclusion if they were ≥65 years old at the time of admission and were hospitalized for an acute burn injury. Burn injuries included thermal, scald, contact, flame, chemical, or electrical mechanisms. Exclusion criteria included readmission for a previously treated burn injury, incomplete documentation of frailty variables or key burn characteristics required for analysis, or loss to follow-up for burn care. A total of 326 unique patients met inclusion criteria and were included in the analysis. After excluding one patient with missing key variables, the final study cohort comprised 325 individuals.

### Data collection

Frailty-related variables were collected through detailed review of geriatric medicine consultations, admission documentation, and functional assessments recorded in the EMR. Frailty factors included fall history, defined as one or more falls within the year preceding the burn injury, and baseline functional status assessed using dependence in Activities of Daily Living (ADLs) and Instrumental Activities of Daily Living (IADLs).

ADLs were defined as basic self-care tasks essential for personal functioning (bathing, dressing, toileting, continence, feeding, and self-transfers), whereas IADLs were defined as higher-order tasks required for independent living (managing medications, housekeeping, meal preparation, shopping, using the telephone, transportation, and financial management). Although fall history was collected as a component of frailty assessment, it was not included in the final multivariable models due to limited completeness.

Functional dependence was categorized as no dependence (no assistance required), some dependence (assistance required for at least one ADL or IADL), or full dependence (assistance required for all ADLs or IADLs). Burn-related variables abstracted from the institutional burn registry included TBSA burn, burn location by anatomical region, 30-day readmission status, presence of inhalation injury confirmed by bronchoscopy or clinical documentation, and renal insufficiency. Renal insufficiency was defined as the presence of acute kidney injury or chronic kidney disease, an estimated glomerular filtration rate (eGFR) <60 mL/min/1.73 m², or elevated serum creatinine (>1.5 mg/dL in men and >1.2 mg/dL in women). Comorbid conditions were obtained from the burn registry, verified by chart review, and were categorized by organ system into cardiovascular, respiratory, and endocrine or metabolic disease (Supplemental Table 1).

### Outcome measures

The primary outcomes were in-hospital mortality and discharge disposition. In-hospital mortality was defined as survival to discharge versus death during the index hospitalization.

Discharge disposition was categorized as discharge to home or home health services versus discharge to a post-acute care facility, including skilled nursing facilities, inpatient rehabilitation units, long term care, and hospice. Thirty-day readmission and thirty-day post-discharge mortality were initially considered but excluded from analysis due to the small number of events. For disposition analyses, patients who died during the index hospitalization or left against medical advice were excluded.

### Statistical analysis

Descriptive statistical analysis was used to summarize demographic and clinical characteristics. Categorical variables were compared using chi-square tests. Associations between clinical variables and study outcomes were assessed using logistic regression for in-hospital mortality and discharge disposition and linear regression for length of stay (LOS).

Variables with *p* <0.10 in bivariate analyses were entered into multivariable models. Model fit and explained variance were evaluated using Nagelkerke R² for logistic models and adjusted R² for linear models. Odds ratios (ORs) and 95% confidence intervals (CIs) were reported.

Multicollinearity was assessed prior to multivariable modeling. All analyses were performed using SPSS Statistics, with *p* <0.05 considered statistically significant.

### Ethical considerations

The study was approved by the Virginia Commonwealth University Institutional Review Board with a waiver of informed consent due to the retrospective nature of the analysis.

## RESULTS

### Study population

A total of 326 geriatric patients with burn injuries met inclusion criteria for this retrospective analysis. One patient was excluded from the analysis due to missing data, resulting in a final analytical sample of 325 patients. With regards to burn-specific factors, the median TBSA burn was 2.0% [IQR, 0.5–7.0%] (Table 1). Approximately 38.5% (N=125) of patients had inhalation injury and 40.9% (N=133) had renal insufficiency. The most common burn locations were the head, face, and neck (N=110; 33.8%) and lower extremities(N=92; 28.3%).

**Table 1.** Characteristics of the Study Population by Burn-Specific Factors, Burn Location, Comorbidity, Functional Status, Hospital Outcomes, and Disposition.

| Characteristics | Frequency or Median |
| --- | --- |
| **Burn Specific Factors** |  |
| Total burn surface area, median [IQR] (%) | 2.0 [0.5–7.0] |
| Inhalation injury, n (%) | 125 (38.5) |
| Renal insufficiency, n (%) | 133 (40.9) |
| **Burn Location<sup>1</sup>** |  |
| Head, face, neck, n (%) | 110 (33.8) |
| Trunk, n (%) | 89 (27.4) |
| Arm, n (%) | 87 (26.8) |
| Hand, n (%) | 79 (24.3) |
| Pelvis, n (%) | 11 (3.4) |
| Leg, n (%) | 92 (28.3) |
| Foot, n (%) | 42 (12.9) |
| **Comorbidity<sup>1</sup>** |  |
| Cardiovascular, n (%) | 254 (78.1) |
| Respiratory, n (%) | 101 (31.1) |
| Metabolic/endocrine, n (%) | 139 (42.8) |
| **Total Organ System** |  |
| Zero, n (%) | 39 (12.0) |
| One, n (%) | 110 (33.8) |
| Two, n (%) | 146 (44.9) |
| Three, n (%) | 31 (9.5) |

| **ADL Status** |  |
| --- | --- |
| Unknown ADL, n (%) | 28 (8.6) |
| No ADL dependence, n (%) | 246 (82.8) |
| Some ADL dependence, n (%) | 37 (12.5) |
| Full ADL dependence, n (%) | 14 (4.7) |
| **IADL Status** |  |
| Unknown IADL, n (%) | 28 (8.6) |
| No IADL dependence, n (%) | 170 (57.2) |
| Some IADL dependence, n (%) | 58 (19.5) |
| Full IADL dependence, n (%) | 70 (23.6) |
| **Hospital Outcomes** |  |
| Length of stay, median [IQR] (days) | 8 [3-16] |
| Alive at discharge, n (%) | 289 (88.9) |
| In-hospital mortality, n (%) | 36 (11.1) |
| AMA, n (%) | 4 (1.2) |
| 30-day readmissions, n (%) | 8 (2.5) |
| 30-day mortality, n (%) | 3 (0.9) |
| **Disposition** |  |
| Home/home health care, n (%) | 195 (60.0) |
| Post-acute care, n (%) | 90 (27.7) |

Abbreviations: ADL=activities of daily living; IADL=instrumental activities of daily living;
<sup>1</sup>The percentages may not sum to 100% since patients may have multiple comorbidities and burn locations

With regards to comorbidities, approximately 78.1% (N=254) of patients had cardiovascular conditions, 42.8% (N=139) had metabolic or endocrine conditions, and 31.1% (N=101) had respiratory conditions (Table 1, Supplemental Table 1). Most patients had comorbidities affecting two organ systems (N=146; 44.9%). Pre-injury functional status was categorized using ADLs and IADLs. ADL status was available for 297 patients and was distributed as follows: 246 patients (82.8%) had no ADL dependence, 37 patients (12.5%) had some ADL dependence, and 14 patients (4.7%) had full ADL dependence prior to injury. IADL status was available for 297 patients with 170 patients (57.2%) with no IADL dependence, 58 patients (19.5%) with some IADL dependence, and 70 patients (23.6%) with full IADL dependence. 28 patients (8.6%) had missing ADLs and IADLs data due to lack of documentation during EMR review. No systematic pattern related to patient characteristics was observed. These patients were excluded from the analyses involving ADL/IADL variables but were retained in models assessing other predictors.

Regarding hospital outcomes, the median hospital LOS was 8 days [IQR, 3–16 days] (Table 1). In-hospital mortality occurred in 11.1% (N=36) of patients during the study period. Among survivors, 1.2% (N=4) were discharged against medical advice (AMA), 60% (N=195) were discharged home or with home health services, and 27.7% (N=90) were discharged to post-acute care facilities. Approximately 2.5% (N=8) of patients experienced 30-day readmission and 0.9% (N=3) had 30-day mortality; given these small sample sizes, we did not conduct descriptive analyses or regressions for these two outcomes.

### In-hospital mortality

Pre-injury ADL dependence demonstrated a significant unadjusted association with in-hospital mortality (*p*=0.002), with mortality risk increasing with worse functional dependence. Among patients with no ADL dependence, 3.3% (N=8/246) died during hospitalization (Table 2). In contrast, mortality was moderately higher among patients with some ADL dependence (16.2%, N=6/37) and among those with full ADL dependence (14.3%, N=2/14).

**Table 2.** Unadjusted Hospital Outcomes and Discharge Disposition by ADL Status.

| **ADL Status** | **Mortality Rate, n (%)** | **AMA Rate, n (%)** | **Post-Acute Care Placement Rate<sup>1</sup>, n (%)** |
| --- | --- | --- | --- |
| No ADL dependence | 8/246 (3.3) | 3/238 (1.3) | 73/235 (31.1) |
| Some ADL dependence | 6/37 (16.2) | 1/31 (3.2) | 11/30 (36.7) |
| Full ADL dependence | 2/14 (14.3) | 0/12 (0.0) | 4/12 (33.3) |

Abbreviations: ADL=activities of daily living; AMA=against medical advice
<sup>1</sup>Post-acute care placement rates are calculated among patients discharged alive with available disposition data; patients who expired in-hospital or were discharged AMA were excluded from disposition analyses

### Model 1A

In a multivariable logistic regression model including ADL status and TBSA burn as independent variables, the overall model was statistically significant (*p*<0.001) and explained 18.4% of the variance in mortality (R² = 0.184). After adjustment for TBSA burn, ADL dependence remained an independent predictor of mortality (*p*=0.004). Compared with patients with no ADL dependence, those with some ADL dependence had a 6.22-fold higher odds of in-hospital mortality (OR, 6.22; 95% CI, 1.87–20.99; *p*=0.003) and those with full ADL dependence had 7.60-fold higher odds (OR, 7.60; 95% CI, 1.36–42.30; *p*=0.021) (Table 3). After adjustment for ADL status, TBSA percentage was also independently associated with in-hospital mortality, wherein each additional 1% increase in TBSA was associated with a 7% increase in the odds of in-hospital mortality (OR, 1.07; 95% CI, 1.03–1.12; *p*=0.001).

**Table 3.** Multivariable Logistic Regression Models for In-Hospital Mortality.

| Predictor | Model 1A |  | Model 1B |  | Model 1C |  |
| --- | --- | --- | --- | --- | --- | --- |
|  | aOR (95% CI) | P value | aOR (95% CI) | P value | aOR (95% CI) | P value |
| **ADL Dependence** |  |  |  |  |  |  |
| No | 1.00 (reference) | – | 1.00 (reference) | – | 1.00 (reference) | – |
| Some | **6.22 (1.87–20.99)** | **.003** | **4.99 (1.44–17.30)** | **.011** | 20.04 (0.75–533.40) | .076 |
| Full | **7.60 (1.36–42.30)** | **.021** | 4.75 (0.81–27.79) | .084 | 4.17 (0.11–160.33) | .435 |
| **IADL Dependence** |  |  |  |  |  |  |
| No | – | – | – | – | 1.00 (reference) | – |
| Some | – | – | – | – | 0.11 (0.003–3.84) | .240 |
| Full | – | – | – | – | 0.25 (0.01–4.34) | .335 |
| **Burn Specific Factors** |  |  |  |  |  |  |
| Total burn surface area | **1.07 (1.03–1.12)** | **.001** | **1.07 (1.02–1.11)** | **.006** | 1.09 (1.00–1.20) | .063 |
| Inhalation injury | – | – | **6.12 (1.63–22.99)** | **.007** | 3.37 (0.42–27.06) | .255 |
| Renal insufficiency | – | – | – | – | **9.611 (1.22–75.56)** | **.031** |
| **Comorbidity<sup>1</sup>** |  |  |  |  |  |  |
| Respiratory comorbidity | – | – | – | – | **180.73 (3.52–9289.13)** | **.010** |
| Cardiovascular | – | – | – | – | 5.43 (0.19– | .326 |
| comorbidity |  |  |  |  | 156.27) |  |
| **Burn location** |  |  |  |  |  |  |
| Head, face, neck | – | – | – | – | 2.67 (0.38–18.64) | .326 |
| Trunk | – | – | – | – | 0.24 (0.02–3.07) | .307 |
| Arm | – | – | – | – | 2.65 (0.29–24.46) | .391 |
| Hand | – | – | – | – | 1.92 (0.29–12.60) | .498 |
| Pelvis | – | – | – | – | 1.76 (0.06–51.63) | .747 |
| Leg | – | – | – | – | 0.44 (0.05–3.96) | .472 |
| Foot | – | – | – | – | 5.85 (0.59–57.93) | .132 |

Abbreviations: aOR=adjusted odds ratio; ADL=activities of daily living; IADL=instrumental activities of daily living
<sup>1</sup>Metabolic/endocrine comorbidity excluded due to collinearity

### Model 1B

When inhalation injury was added to the previous model as a third independent variable, the overall model fit improved (R² = 0.264). In this expanded model, some ADL dependence remained significantly associated with in-hospital mortality (*p*=0.011) (Table 3). After adjustment for both TBSA burn and inhalation injury, patients with some ADL dependence had 4.99-fold higher odds of in-hospital mortality compared with those with no ADL dependence (OR = 4.99; 95% CI, 1.44–17.30; *p*=0.011), whereas full ADL dependence was no longer significantly associated with mortality (*p*=0.084). After adjustment for both ADL status and inhalation injury, TBSA burn percentage remained an independent predictor of in-hospital mortality, wherein each additional 1% increase in TBSA was associated with a 7% increase in mortality odds (OR, 1.07; 95% CI, 1.02–1.11; *p*=0.006). Finally, after adjusting for ADL status and TBSA burn, inhalation injury was significantly associated with mortality, exhibiting a 6.12-fold increase in the odds of in-hospital mortality (OR, 6.12; 95% CI, 1.63–22.99; *p*=0.007).

### Model 1C

A separate multivariable model including all covariates demonstrated moderate explanatory performance for in-hospital mortality, accounting for 54.9% of the variance in the hospital outcome (R² = 0.549). Within this model, respiratory comorbidity emerged as the strongest independent predictor of mortality, with affected patients experiencing a 180.73-fold increase in the odds of in-hospital mortality (OR, 180.73; 95% CI, 3.52–9289.13; *p*=0.010); however, the wide confidence interval indicates this estimate should be interpreted with caution (Table 3). Renal insufficiency was also independently associated with mortality, with a 9.61-fold higher odds of death (OR, 9.611; 95% CI, 1.22–75.56, *p*=0.031). Other covariates, including TBSA, inhalation injury, cardiovascular comorbidity, burn locations, and functional status, were not independently associated with in-hospital mortality in this fully adjusted model.

### Discharge disposition

Discharge disposition analyses were limited to patients discharged alive and excluded those discharged AMA. A total of four patients were discharged AMA, including three patients with no ADL dependence and one patient with some ADL dependence (Table 2). In unadjusted analyses, ADL status was not a significant predictor of discharge disposition (*p*=0.819). Among the 285 patients who were discharged alive and did not leave AMA, 90 patients were discharged to post-acute care (Table 1). Rates of post-acute care placement were similar across ADL status, occurring in 31.1% (N=73/235) of patients with no ADL dependence, 36.7% (N=11/30) of those with some ADL dependence, and 33.3% (N=4/12) of those with full ADL dependence (Table 2).

### Model 2A

In a multivariable logistic regression model including ADL status and TBSA burn as independent variables, the model was significant (*p*<0.001) and explained a modest proportion of variance in discharge disposition (R² = 0.141). After adjustment for TBSA, ADL status was not a significant predictor of discharge disposition (*p*=0.462) (Table 4). After adjustment for ADL status, TBSA percentage emerged as the primary predictor of post-acute care placement, wherein each additional 1% increase in TBSA was associated with a 12.2% increase in the odds of post-acute care discharge (OR, 1.12; 95% CI, 1.07–1.18; *p*<0.001).

**Table 4.** Multivariable Logistic Regression Models for Discharge Disposition.

| Predictor | Model 2A<sup>1,2</sup> |  | Model 2B<sup>1,2</sup> |  | Model 2C<sup>1,2</sup> |  |
| --- | --- | --- | --- | --- | --- | --- |
|  | aOR (95% CI)<sup>2</sup> | P value | aOR (95% CI)<sup>3</sup> | P value | aOR (95% CI)<sup>4</sup> | P value |
| **ADL Dependence** |  |  |  |  |  |  |
| No | 1.00 (reference) | – | 1.00 (reference) | – | 1.00 (reference) | – |
| Some | 1.51 (0.68–3.32) | .309 | 1.47 (0.66–3.25) | .344 | 1.04 (0.35–3.09) | .950 |
| Full | 3.20 (0.92–11.07) | .067 | 2.81 (0.81–9.80) | .105 | 0.86 (0.18–4.17) | .850 |
| **IADL Dependence** |  |  |  |  |  |  |
| No | – | – | – | – | 1.00 (reference) | – |
| Some | – | – | – | – | 2.48 (1.05–5.90) | .039 |
| Full | – | – | – | – | 2.61 (1.13–6.05) | .025 |
| **Burn Specific Factors** |  |  |  |  |  |  |
| Total burn surface area | 1.12 (1.07–1.18) | <.001 | 1.14 (1.08–1.20) | <.001 | 1.19 (1.08–1.30) | <.001 |
| Inhalation injury | – | – | 2.09 (1.17–3.75) | .013 | 2.95 (1.40–6.22) | .004 |
| Renal insufficiency | – | – | – | – | 2.75 (1.45–5.20) | .002 |
| **Comorbidity<sup>2</sup>** |  |  |  |  |  |  |
| Respiratory comorbidity | – | – | – | – | 0.38 (0.15–0.99) | .047 |
| Cardiovascular comorbidity | – | – | – | – | 0.50 (0.16–1.56) | .225 |

| Burn location |  |  |  |  |  |  |
| --- | --- | --- | --- | --- | --- | --- |
| Head, face, neck | – | – | – | – | **0.45 (0.20–0.99)** | **.048** |
| Trunk | – | – | – | – | 0.78 (0.32–1.88) | .584 |
| Arm | – | – | – | – | 0.62 (0.26–1.49) | .272 |
| Hand | – | – | – | – | 0.86 (0.37–2.00) | .729 |
| Pelvis | – | – | – | – | **16.54 (1.39–196.66)** | **.026** |
| Leg | – | – | – | – | 0.80 (0.36–1.77) | .578 |
| Foot | – | – | – | – | **2.46 (1.00–6.04)** | **.049** |

Abbreviations: aOR=adjusted odds ratio; ADL=activities of daily living; IADL=instrumental activities of daily living; AMA=against medical advice
<sup>1</sup>Analysis restricted to patients discharged alive and not AMA
<sup>2</sup>Bolded numbers represent data of statistical significance
<sup>3</sup>Metabolic/endocrine comorbidity excluded due to collinearity

### Model 2B

When inhalation injury was added to the previous model as a third independent variable, overall model performance improved slightly (R² = 0.168). After adjustment for TBSA and inhalation injury, ADL status again did not demonstrate a significant association with discharge disposition (*p*=0.585) (Table 4). After adjustment for ADL status and inhalation injury, TBSA percentage remained an independent predictor of post-acute care placement, with each additional 1% increase in TBSA associated with a 13.6% increase in odds (OR, 1.14; 95% CI, 1.08–1.20; *p*<0.001). After adjustment for ADL status and TBSA, inhalation injury was also a significant predictor, showing patients with inhalation injury having 2.09-fold higher odds of post-acute care placement (OR, 2.09; 95% CI, 1.17–3.75; *p*=0.013).

### Model 2C

A separate model including all covariates demonstrated moderate explanatory performance, accounting for 48.1% of the variance in discharge disposition (R² = 0.481). Variables associated with higher odds of post-acute care placement included TBSA percentage (OR, 1.19; 95% CI, 1.08–1.30; *p*<0.001), inhalation injury (OR, 2.95; 95% CI, 1.40–6.22; *p*=0.004), renal insufficiency (OR, 2.75; 95% CI, 1.45–5.20; *p*=0.002), pelvic burns (OR, 16.54; 95% CI, 1.39–196.66; *p*=0.026), foot burns (OR, 2.46; 95% CI, 1.00–6.04, *p*=0.049) and both some (OR, 2.48; 95% CI, 1.05–5.90; *p*=0.039) and full (OR, 2.61; 95% CI, 1.13–6.05; *p*=0.025)

IADL dependence (Table 4). In contrast, respiratory comorbidity (OR, 0.38; 95% CI, 0.15–0.99; *p*=0.047) and head, face, neck burns (OR, 0.45; 95% CI 0.20–0.99; *p*=0.048) were associated with lower odds of post-acute care placement. Cardiovascular comorbidity and ADL status were not independently associated with post-acute care placement in this fully adjusted model.

## DISCUSSION

In this cohort of geriatric patients with burn injuries, we evaluated pre-injury functional status, burn-specific factors, comorbidity burden, and burn location as prognostic factors for in-hospital mortality and post-acute care disposition. Building on prior literature demonstrating that frailty in older adults is associated with mortality and discharge outcomes, this study contributes a novel operationalization of frailty by examining ADL and IADL dependence separately. Our findings demonstrate that ADL dependence is independently associated with higher odds of in-hospital mortality, whereas IADL dependence is associated with increased odds of post-acute care disposition. In the fully adjusted models, burn-specific factors (TBSA and inhalation injury) and specific burn locations (pelvis, foot, head, face, or neck) were associated with post-acute care placement, but not mortality. Collectively, these findings indicate that functional dependence captures clinically meaningful vulnerability in older burn patients that is not fully reflected by traditional burn severity metrics alone, and that different domains of functional impairment may influence distinct clinical outcomes.

Frailty has increasingly been recognized as a key predictor of outcomes in older adults with burn injuries; however, which domains of frailty are most clinically informative in burn care remains understudied. Most existing studies have relied on global frailty measures, such as the CFS, to demonstrate associations with mortality, complications, and discharge disposition, without examining how individual components of frailty differentially contribute to these outcomes.^8–10^ While these studies establish frailty as a meaningful prognostic construct, they do not clarify which specific elements of frailty influence distinct outcomes or how frailty interacts with traditional burn severity metrics.

Our study separately operationalizes pre-injury functional dependence into ADLs and IADLs and examines their differential associations with in-hospital mortality and discharge disposition. ADL dependence remained significantly associated with in-hospital mortality after adjusting for TBSA and inhalation injury, with patients exhibiting some ADL dependence experiencing nearly five-fold higher odds of death during hospitalization. This finding supports prior work demonstrating that frailty is associated with increased mortality in older burn patients independent of chronological age, TBSA, and inhalation injury, supporting the idea that diminished physiologic reserve contributes to poor outcomes in this population.^8–10,12^ In contrast, IADL dependence was not significantly associated with in-hospital mortality in the fully adjusted model.

A different pattern emerged when examining discharge disposition. Whereas ADL dependence was not independently associated with discharge disposition, IADL dependence emerged as a significant predictor of post-acute care placement in the fully adjusted model. Both some and full IADL dependence were associated with more than two-fold higher odds of discharge to post-acute facilities. IADLs reflect higher-order functional capabilities, such as medication management, transportation, and household tasks, which are critical for safe independent living after hospital discharge. This finding is consistent with prior evidence showing that IADL impairment is independently associated with an increased risk of nursing home admission. For example, one study of middle-aged adults reported a 73% higher hazard among individuals with IADL impairment, while another study of geriatric adults found a 31% higher hazard.^9,11^

In the context of burn patients, while prior literature has identified associations between frailty and institutional discharge, most studies do not differentiate between ADL and IADL domains. Our findings suggest that ADLs and IADLs capture distinct aspects of vulnerability, with ADLs more closely related to physiologic reserve and survival, and IADLs more relevant to post-hospital functional needs and discharge disposition. However, one possible explanation for the lack of association between ADL dependence and discharge disposition is that patients who were already ADL-dependent prior to admission may have had established caregiving support in the home setting, allowing return to their baseline living environment after hospitalization. While caregiver availability was not directly measured in this study, this hypothesis may explain why baseline ADL dependence was less predictive of discharge disposition compared with IADL dependence.

Burn severity, measured by TBSA, and inhalation injury demonstrated significant association with both in-hospital mortality and discharge disposition in partially adjusted models. Higher TBSA was independently associated with increased odds of in-hospital mortality and post-acute care placement. Similarly, inhalation injury significantly increased the odds of both adverse outcomes, which may reflect the added physiologic burden and respiratory compromise associated with airway involvement. These findings are consistent with extensive prior literature identifying burn size and inhalation injury as central determinants of adverse outcomes in older burn patients.^14^ Collectively, our results reinforce that TBSA and inhalation injury remain important prognostic factors and should continue to be prioritized in the development and refinement of prognostic models for geriatric burn populations.

With respect to comorbidities, respiratory comorbidity and renal insufficiency were independently associated with higher odds of in-hospital mortality. Prior evidence corroborates this finding, identifying chronic obstructive pulmonary disease (COPD) and renal insufficiency as major contributors to mortality in geriatric burn populations, with one study suggesting a 56% higher risk of survival among burn patients with respiratory comorbidity compared to those without.^15,16^

In contrast, pre-existing respiratory comorbidity was associated with significantly lower odds of post-acute care placement. In the context of elevated in-hospital mortality, a plausible mechanism for this finding could be selection bias due to differential survival, wherein patients with respiratory comorbidity who survive to discharge represent a subset with less severe burns or greater baseline physiologic reserve. Supporting this interpretation, one study showed that burn patients with COPD are less likely to require surgical intervention compared to those without COPD.^15^ To date, few studies have examined the relationship between respiratory comorbidity and discharge disposition, warranting further investigation.

Burn injuries involving the pelvis, foot, head, face, or neck were significantly associated with discharge disposition. Our study findings show that burns involving the pelvis or foot were both associated with higher odds of post-acute care placement. This is consistent with prior literature showing that pelvic burns were associated with a two-fold higher risk of receiving surgical interventions, contributing to worse prognosis and complications.^17^ Similarly, foot burns have been identified as independent predictors of prolonged inpatient rehabilitation, which may reflect mobility limitations and wound-healing challenges involving weight-bearing structures.^18^ In contrast, our study found that burns involving the head, face, or neck were associated with lower odds of post-acute care placement among survivors.

Prior literature presents mixed evidence regarding this relationship. Some studies suggest that severe head and neck burns, particularly full-thickness injuries or those accompanied by inhalation injury, are associated with increased mortality, greater intensive care utilization, higher likelihood of transfer to specialized burn centers, and increased post-acute care needs.^19–22^ However, another study reported that facial burns were associated with a 23% lower odds of readmission compared with burns at other anatomical sites.^23^ In contrast, that same study found that third-degree burns were associated with higher readmission rates. Taken together, these findings suggest that while initial hospitalization for facial burns may be complex, isolated facial burns without extensive severity or complications may be associated with lower post-acute care requirements. Further research is needed to clarify how burn severity moderates the relationship between burn location and post-acute care disposition.

Overall, these findings suggest that early assessment of pre-injury ADL dependence may help identify geriatric burn patients at elevated risk for in-hospital mortality, even among those with relatively small TBSA injuries, and may support earlier goals-of-care discussions, closer physiologic monitoring, or proactive geriatric and palliative care consultation. Assessment of IADL dependence may be particularly useful for anticipating discharge needs and facilitating early involvement of rehabilitation services, case management, and social work to streamline transitions to post-acute care. Incorporating simple, routinely collected functional measures into burn assessment may therefore improve prognostication, enhance risk stratification, and promote more individualized, goal-oriented care.

### Limitations

This study has several limitations. First, the retrospective, single-center design precludes causal inference and may limit generalizability to other burn centers with different patient populations, care practices, or discharge resources. Second, although the overall sample size was moderate, the number of patients with full ADL dependence and certain burn locations was relatively small, resulting in wide confidence intervals for some estimates, particularly in fully adjusted models. Third, pre-injury functional status was abstracted from clinical documentation rather than assessed using standardized, prospective instruments, introducing the potential for misclassification or documentation bias. Fourth, while we adjusted for several burn severity metrics and comorbidity burden, there may still be residual confounding due to unmeasured factors (e.g., social support, baseline cognitive impairment, other frailty domains, and post-discharge resource availability) that influence discharge disposition. Fifth, discharge disposition was used as a measure for post-hospital functional needs; however, placement decisions are also influenced by institutional practices, insurance status, and availability of post-acute care resources, which were not captured in this analysis. Finally, the small number of 30-day readmission and mortality events precluded meaningful analysis of longer-term outcomes, limiting insight into post-discharge recovery trajectories.

## CONCLUSION

In this retrospective cohort of geriatric patients with burn injuries, pre-injury functional status demonstrated outcome-specific prognostic value. ADL dependence was independently associated with in-hospital mortality, whereas IADL dependence was associated with post-acute care placement, highlighting the importance of distinguishing between basic and instrumental functional limitations in older burn patients. Traditional burn severity measures, including TBSA and inhalation injury, remained key determinants of both mortality and discharge disposition, while comorbidity burden and anatomic burn location further shaped outcome trajectories.

Together, these findings highlight the importance of incorporating structured functional assessment with established burn severity metrics to improve prognostication, inform clinical decision-making, and support individualized care for older adults with burn injuries.

## Data Availability

All data produced in the present work are contained in the manuscript.

