## Supplemental Table 1 for "Frailty-based prognostication of clinical outcomes in geriatric burn patients: a retrospective study"

**Supplemental Table 1. Clinical Conditions by Comorbidity Category**

| **Comorbidity Category** | **Clinical Conditions** |
| --- | --- |
| Cardiovascular | - Abdominal aortic aneurysm - Arrhythmia - Cardiac history - Congestive heart failure - Coronary artery disease - Hyperlipidemia - Hypertension - Myocardial infarction - Peripheral arterial disease |
| Respiratory | - Asthma - Chronic obstructive pulmonary disease - Emphysema - Respiratory failure |
| Metabolic/Endocrine | - Diabetes mellitus - Hypothyroidism - Obesity |
